# Endoscopic ultrasound-guided biliary rendezvous after failed cannulation, and comparison between benign vs malignant biliopancreatic disorders: outcomes at a single tertiary-care center

**DOI:** 10.1101/2024.05.09.24307139

**Authors:** Joan B Gornals, Albert Sumalla-Garcia, Sergi Quintana, Daniel Luna-Rodriguez, Julio G Velasquez-Rodriguez, Maria Puigcerver-Mas, Julia Escuer, Sandra Maisterra, Mar Marin, Virginia Munoa, Berta Laquente, Juli Busquets

**Author notes:** No financial disclosures. No grant support. All authors have read and agreed on the content of this manuscript. **Author contributions as follows:** JBG conceived the project and designed the study. JBG, AGS, and SQ provided a critical review and participated in the design of the study. MM, BL, JB, and AGS promoted the enrolment of patients. DL, JGV, SM, SQ, MP, JE, AGS, and JBG carried out acquisition, analysis, and interpretation of data. VM, AGS, and JBG did the statistical analysis. AGS and JBG drafted the manuscript, interpreted the data, verified the underlying data, and critically reviewed the manuscript. JBG had final responsibility for the decision to submit for publication. All authors read, revised, and provided a critical review of the draft manuscript. All authors approved the final manuscript. **Corresponding author:** Joan B. Gornals, M.D, PhD. Endoscopy Unit, Dept. of Digestive Diseases, Hospital Universitari de Bellvitge; IDIBELL (Bellvitge Biomedical Research Institute). Associate Professor, Universitat de Barcelona, Feixa Llarga s/n 08907 L’Hospitalet de Llobregat, Barcelona, Catalonia, Spain.

## Abstract

**Background:** Endoscopic ultrasound (EUS)-guided biliary rendezvous (RV) is an EUS-assisted technique described as a rescue method in cases of failed biliary cannulation via endoscopic retrograde cholangiography (ERC). Current literature remains unclear regarding its current role. The study aim was to evaluate the effectiveness for biliary EUS-RV, and comparison between benign vs malignant biliopancreatic disorders.

**Methods:** Retrospective observational study with prospective consecutive inclusion in a specific database from a tertiary-center. All patients with biliopancreatic diseases that underwent a EUS-assisted ERC between October-2010 and November-2022 for failed ERC were included. Main outcomes were technical/overall success. Secondary outcomes were safety, potential factors related to failure/success or safety; and a comparative analysis between EUS-RV and EUS-guided transmural drainage (TMD) in malignant cases.

**Results:** A total of 69 patients who underwent EUS-RV procedures, with benign and malignant pathologies (n=40 vs n=29), were included. Technical/overall success and related-adverse events (AEs) were 79.7% (95%CI, 68.3-88.4)/74% (95%CI, 61-83.7) and 24% (95%CI, 15.1-36.5), respectively. Failed cases were mainly related with guidewire manipulation. Seven failed RV were successfully rescued by EUS-TMD. On multivariable analysis, EUS-RV and malignant pathology was associated with a greater failure rate (technical success: OR,0.21; 95%CI,0.05-0.72; p=0.017), and higher AEs rate (OR,3.46; 95%CI,1.13-11.5; p=0.034). Also, the EUS-TMD group had greater technical success (OR,16.96; 95%CI,4.69-81.62; p<0.001) and overall success (OR, 3.09; 95%CI,1.18-8-16; p<0.026) with a lower AEs rate (OR,0.30; 95%CI,0.11-0.78; p=0.014) than EUS-RV in malignant disorders.

**Conclusions:** EUS-RV is a demanding technique with better outcomes in benign than in malignant biliopancreatic diseases. Comparison of the EUS-TMD group on malignant disorders showed worse outcomes with EUS-RV. Given these findings, maybe EUS-RV is not the best option for malignant biliopancreatic disorders.

## INTRODUCTION

Endoscopic ultrasound (EUS)-guided assisted bile duct access is an indirect technique performed using two endoscopes. It includes EUS-guided rendezvous (EUS-RV) using guidewire, or even colorant injection (‘wireless’ concept), as assistant techniques for failed endoscopic retrograde cholangiography (ERC). It is considered a technically demanding technique not free from adverse events (AEs). In particular, the technical success rates of EUS-biliary RV ranges between 72% and 96%, with a mean of 84-86% in expert hands, and with 10-34% safety [1,2].

Recently, the European Society of Gastrointestinal Endoscopy (ESGE) guidelines suggested EUS-RV after a second failed ERC in benign biliary disease, and with normal GI anatomy, in high volume centers (weak recommendation, low quality evidence, ESGE2021), but its current role in malignant disease is unclear. Surprisingly, the technical success of the EUS-RV technique in benign disease has been reported to be lower than in malignant disease, and AEs are more likely to occur (27%), owing to limited bile duct dilatation and technical difficulty in accessing [1,2].

In case of failed ERC, EUS-guided bile duct access techniques come in several varieties [3]. EUS-RV, although widely recognized, in real-clinical practice is performed in relatively few high-volume centers, and its standardization is still in the process of development. Also, the related literature is still sparse (i.e., 248 malignant cases reported in the latest ESGE guidelines), with mostly retrospective studies with small sample sizes (range; 13 to 58 cases), and long-term outcomes are limited. These are the main reasons why this procedure is not yet widely used twenty years after the demanding technique was initially reported (first EUS-biliary RV, by Mallery et al in 2004) [4].

This is a review of our experience in EUS-guided assisted ERC over a decade, including long-term analysis. The main aim was to evaluate the effectiveness of EUS-guided assisted RV, with a comparison between benign vs. malignant groups.

## MATERIAL AND METHODS

### Study design

In this retrospective single-center study, an EUS/ERCP database maintained for the period between October 2010 and November 2022 was retrospectively reviewed.

The study was approved by our institutional ethics committee (PR240/12), Comitè Ètic d’Investigació Clínica, Hospital Universitari de Bellvitge) and conducted in accordance with the principles of the Declaration of Helsinki and the guidelines for Good Clinical Practice.

### Participants

All consecutive EUS-guided assisted ERC cases were identified and included in this analysis. A previous ERC attempt was mandatory before attempting EUS-guided bile duct access. Exclusion criteria were transpapillary stent by standard ERC, EUS-guided pancreatic duct intervention, and lack of follow-up information.

Figure1 contains a detailed flowchart.

**Figure 1:**
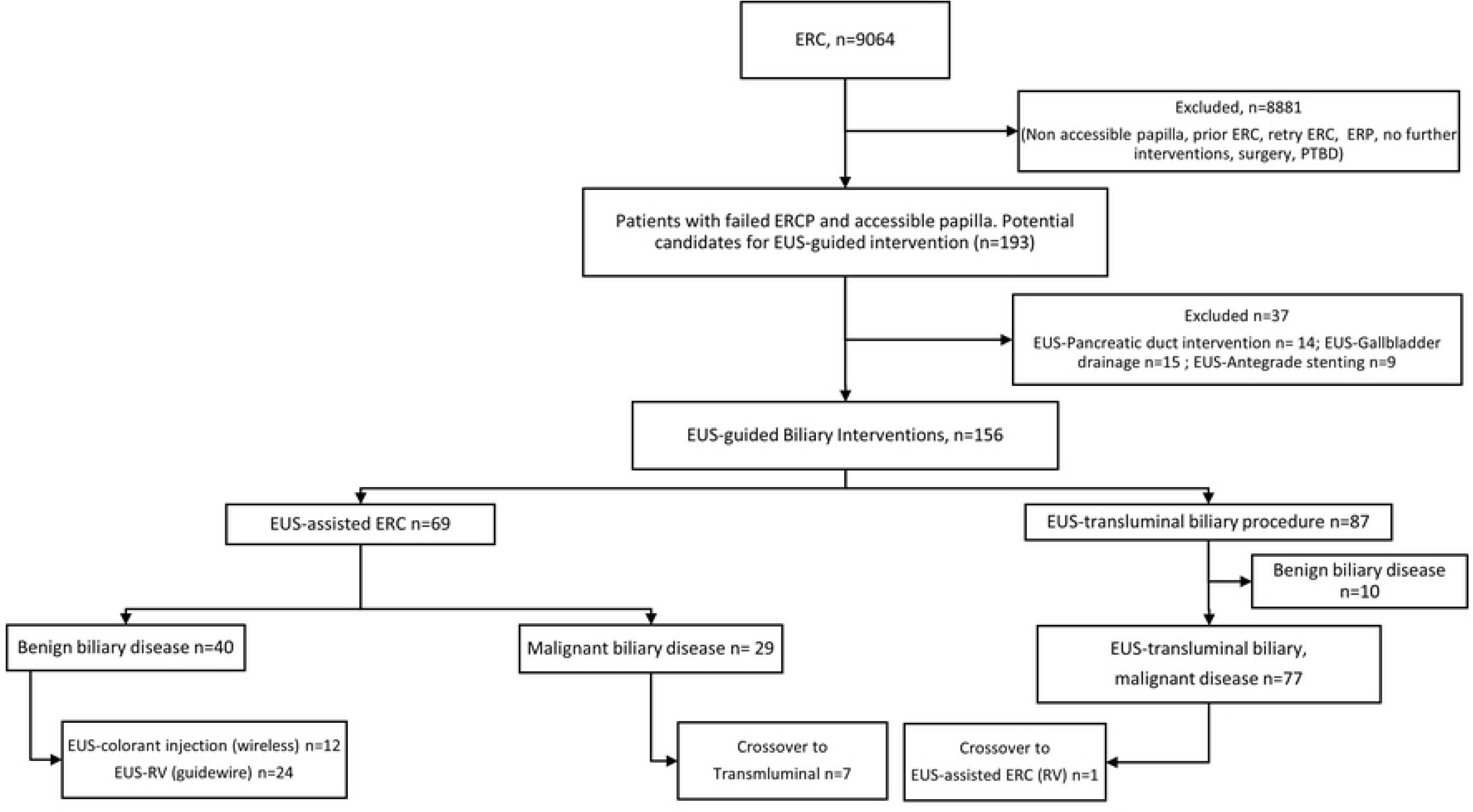
Patient flowchart of the study. EUS-guided interventional biliary procedures.

The following variables were reviewed before and after the interventional procedures: demographic and clinical data, procedure and technical details, follow-up data, incidents, and AEs.

All patients provided written informed consent before each procedure. Data was introduced in a prospectively maintained database, and accessed during all the period of the study. Authors had access to information that could identify individual participants during or after data collection.

### Technique

Orotracheal intubation or deep sedation was provided according to the anesthesiologist’s criteria. All EUS-guided biliary interventions were done by a single endoscopist with expertise in EUS, ERCP, or stenting (>15 years), and with nurses trained in both procedures. Rectal indomethacin and antibiotics were routinely given in all cases. Procedures were performed with patients in the left-side or supine position.

A linear array echoendoscope (GF-UCT140-AL5, GF-UCT180 Olympus; or EG-580UT, Fujifilm) was advanced into the gastric cavity or duodenum and the biliary tree was identified by EUS. Under EUS guidance, the intra- or extrahepatic bile duct was accessed transgastrically/transduodenally, respectively, with a 19-G and/or 22-G fine-aspiration needle and confirmed by bile aspiration. Limited cholangiogram was obtained at the endoscopist’s discretion. Doppler imaging was used to avoid interposal vessels.

All patients were monitored in the recovery room of the endoscopy unit for at least 1 hour and admitted for 24 hours of clinical observation.

#### EUS-guided assisted ERC

i. <u>EUS-guided colorant injection or ‘wireless’ RV</u>: as noted in a previous report [5]. After contrast-medium injection to obtain a cholangiography, a sufficient amount (5-10 mL) of colorant (methylene blue or linoleic acid) was injected, depending on duct diameter and the presence of contrast fluid flow, into the small intestine, and monitored by fluoroscopy. If EUS-guided cholangiography was successful, the echoendoscope was withdrawn and ERC was immediately attempted. Papilla orifice identification was achieved using colorant flow. Once the papilla was reached, a sphincterotome, with a 0.025- or 0.035-inch guidewire, was used for direct cannulation. Secondly, if necessary, a precut with a needle-knife was attempted. If cannulation was not achieved after several attempts (3-4), an EUS-guided RV using guidewire, or a second ERC session, was considered.
ii. <u>EUS-guided RV with guidewire:</u> under fluoroscopic guidance, a guidewire was advanced through the needle and antegradely into the bile duct across the obstruction site to the enteral lumen. When a 19-G EUS needle was used, guidewires were mostly 0.035-inch (jagwire, Boston Sc) or 0.025-inch (Visiglide2, Olympus; or Revolution, angle-tipped, Boston Sc). Using a 22-G EUS-needle, a 0.018-inch needle (Novagold, Boston Sc) was used. Contrast injection was performed at the endoscopist’s discretion.

After guidewire passage through the papilla, echoendoscope was replaced by a duodenoscope thereby allowing conventional ERC to be performed. Generally, retrograde biliary cannulation was attempted alongside the antegrade guidewire using the monorail technique with a homemade modified 3.9/4.4F sphincterotome. In case of failed RV the procedure was finished, or else the guidewire was coiled at the endoscopist’s discretion, and transmural stenting was considered over the wire.

Finally, in both techniques and in accordance with findings, sphincterotomy, retrograde stent placement, or other maneuvers were considered.

Examples of various EUS-RV (benign or malignant disorders; long or short-scope position) are presented in **Figure2 and supplementary figures (FigS1, FigS2, FigS3).**

**Figure 2:**
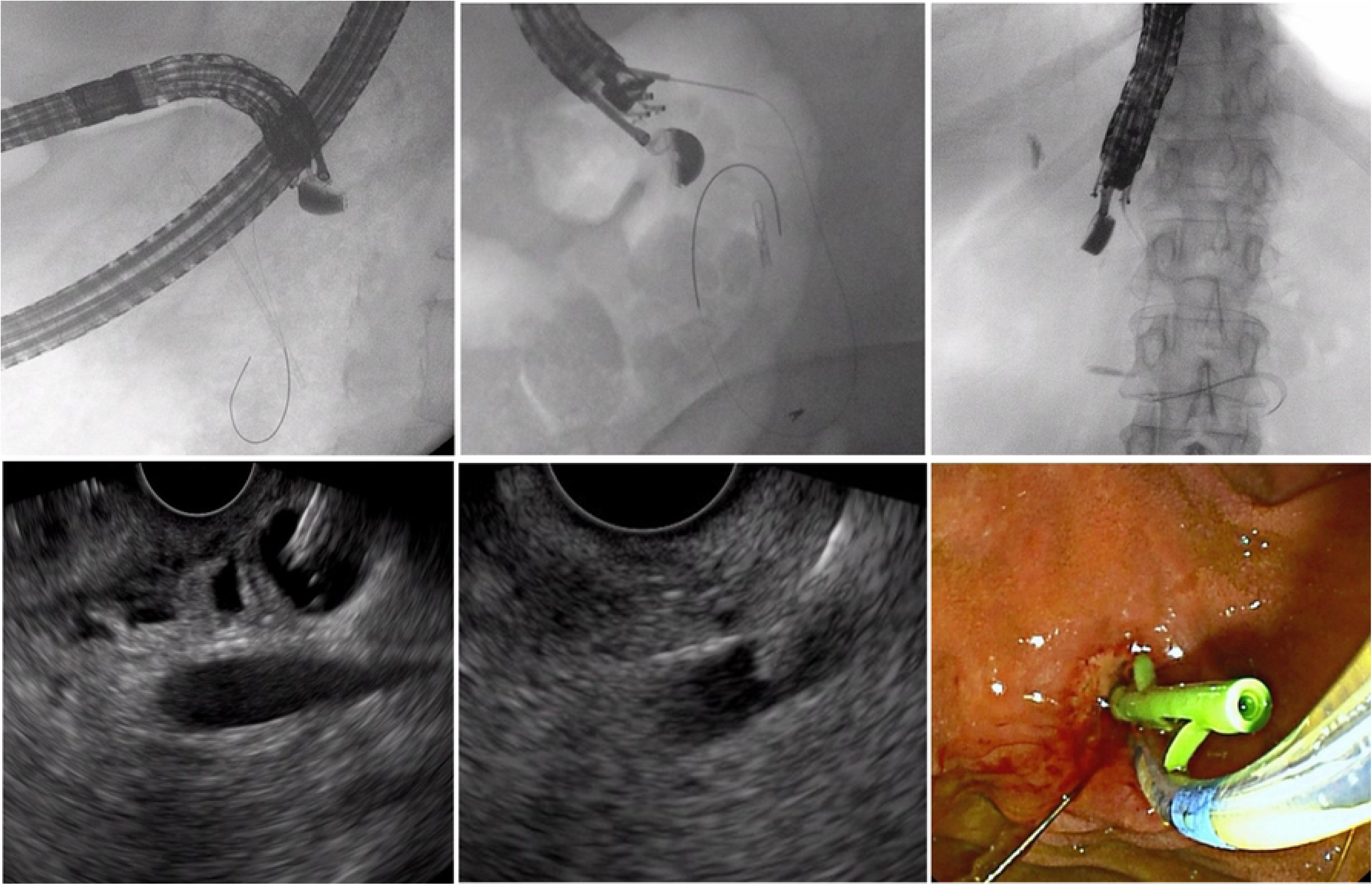
Examples of EUS-guided assisted ERC (Rendezvous guidewire type). **A**, Transduodenal and long-scope position: the extrahepatic bile duct is punctured from the bulb. **B**, Transduodenal short-scope position: the extra-hepatic bile duct is punctured from the bulb. **C**, Transduodenal short-scope position: the extra-hepatic bile duct is punctured from the second portion of the duodenum. **D**, **E**, EUS image during puncture of extrahepatic and intrahepatic bile duct. **F**, Rendezvous with the guidewire passed antegradely through the papilla.

#### EUS-guided biliary TMD

In transluminal cases (e.g. choledochoduodenostomy, hepaticogastrostomy), the entire procedure was performed using a linear echoendoscope. Interventional technique and approach were as detailed in previous published reports [6,7].

With the introduction of biliary LAMS at our unit in 2014, EUS-guided choledochoduodenostomy progressively gained major prominence to become the standard approach in cases of malignant distal biliary obstruction [8]. This change was natural following recognition that the procedure was less time-consuming and with increased confidence with this EUS-biliary TMD variant.

### Follow-up

All laboratory, radiologic, surgical, and clinical findings after index procedures or repeat sessions were reviewed. All imaging parameters were reviewed and taken from the original written reports. The last available clinical follow-up was used to assess patients’ responses to interventions.

### Definitions

<u>Technical success for EUS-RV:</u> defined as successful biliary access with papilla identification or guidewire passage from the biliary system into the small bowel to allow conventional ERC to be performed.

<u>Overall success, RV:</u> included technical success and successful retrograde biliary cannulation.

<u>Technical or clinical success for TMD</u> (according to each clinical indication) or procedure time, defined as previously described [6,7]

<u>Safety</u>: defined as rate of AEs. AEs were recorded for all index and repeat procedures. AE severity was graded according to the AGREE-American Society for Gastrointestinal Endoscopy (ASGE) lexicon for endoscopic AEs.

Failed biliary cannulation with ERC was not standardized.

### Study endpoints

The primary endpoint was to assess the effectiveness of the EUS-guided biliary RV (global and between either benign or malignant pathology), in terms of technical and overall success.

Secondary endpoints were to assess the safety, potential factors related to failure/success or safety; and a comparative analysis between EUS-RV and EUS-TMD for malignant distal biliary obstruction cohort.

### Statistical analysis

The number of cases and percentages were presented as categorical variables; continuous variables were shown as mean and standard deviation (SD), or median and interquartile rank (IQR), depending on whether the data distribution was normal. Categorical variables were compared using the chi-squared test (or Fisher’s exact test if required). Quantitative variables were compared using the student’s *t* test or Kruskall-Wallis test, according to application criteria.

Independent univariate logistic regression models were performed to identify variables associated with success/failure and safety. The models were repeated, adjusted for different clinical, technical, and analytical variables depending on the outcome analysed.

The level of statistical significance was set at <0.05. The statistical package used was R version 4.3.1 for Windows.

## RESULTS

### Demographics and procedure details for EUS-assisted ERC

A total of 69 (43.4% female; mean [SD] age, 73 [9.0] years) EUS-assisted ERC were performed out of 9,072 ERCP (0.7%). Demographic and clinical characteristics are shown in **Table1**.

**Table 1:**
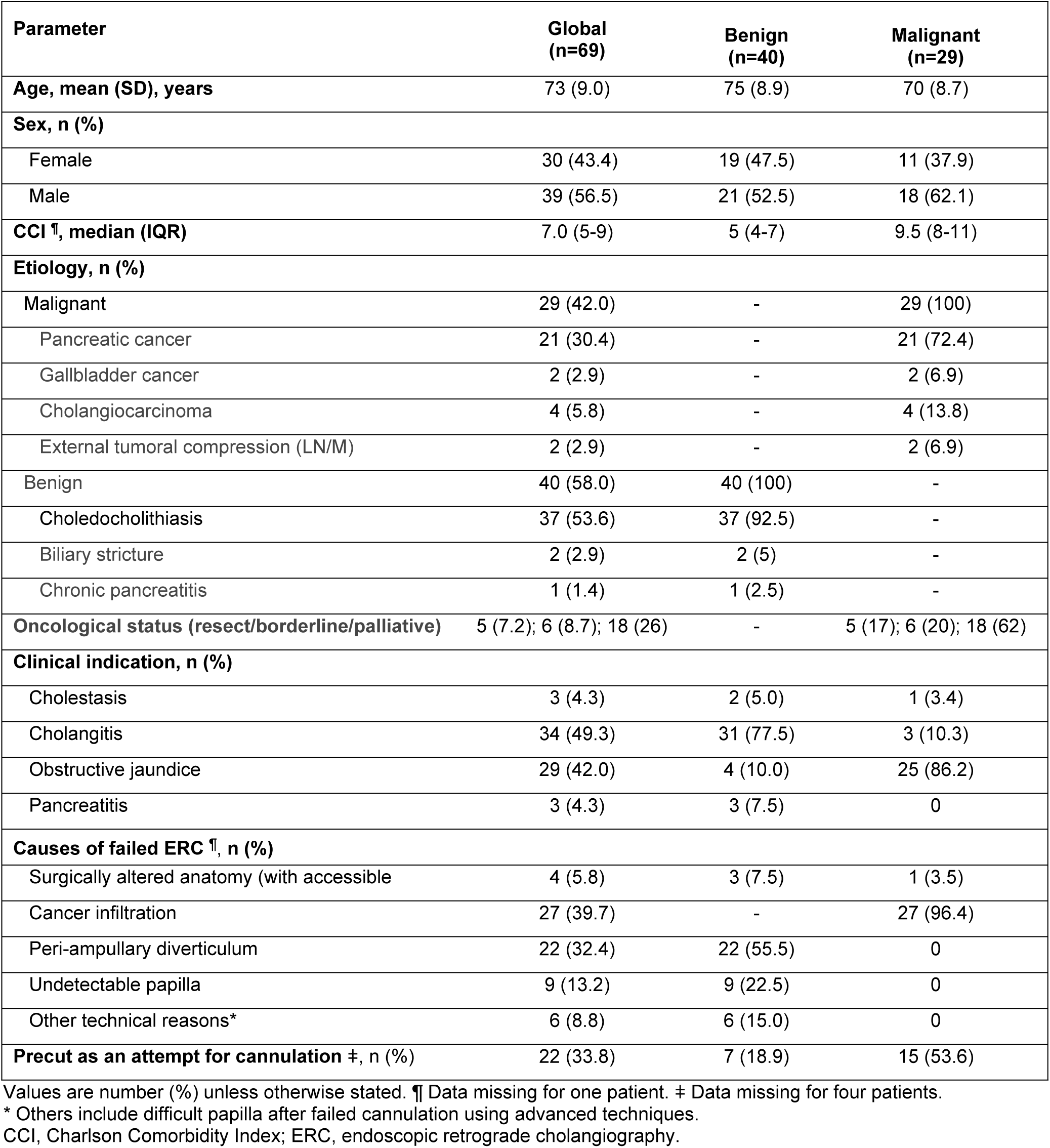
Demographic and clinical data. EUS-guided assisted ERC.

Benign disorders (40 patients [58%]) were more frequent than malignant (29 patients [42%]), with choledocholithiasis (n=37) and pancreatic cancer (n=21) as the most frequent in each category, respectively. The main reason for failed conventional ERC was peri-diverticular papilla (n=22) and tumoral infiltration (n=27) in the benign and malignant groups, respectively.

Concerning EUS-assisted ERC type, most procedures were EUS-RV using guidewire, in 75.4% (n=52), EUS-assisted colorant injection in 17.4% (n=12), and combined techniques in 7.2% (n=5). In all EUS-RV cases in the malignant group, a guidewire was used.

Mean (SD) bile duct diameter and procedure time were 10.8 (3.3) mm and 92 min (IQR,71-110), respectively. A 19G and 22G needle size (n=34 vs. n=31; both needles in 4 cases) were used to access the bile duct in all procedures. Extrahepatic access (82.6%) was more frequently than intrahepatic puncture site (14.5%), and transduodenal with short-scope position (66%) was the most approach most frequently used. EUS-guided assisted ERC was done in the same session, after a failed ERC procedure, in almost half of the cases (47.8%), but in the malignant group this percentage increased to 79.3%.

### Comparison of malignant and benign groups

Significant differences were encountered: wider bile duct diameter (mean (SD) 12.4[2.7] vs 9.7[3.2] mm; p<0.001), longer procedure duration (median, 96 vs 82 mm; p<0.001), shorter time from failed ERC to EUS-RV (2.14 [5.4] vs 18.9 [23] days; p<0.001), greater use of 19G needles (65.5 vs 37.5%; p<0.044), and greater number of EUS-FNA in the same procedure (48.2 vs 20.2%) were detected for the malignant group (**Table2**).

**Table 2:** Procedure details and clinical outcomes of EUS-guided assisted ERC.

| Parameters | Global<br>(n=69) | Benign<br>(n=40) | Malignant<br>(n=29) | p |
| --- | --- | --- | --- | --- |
| **Procedure details:** |  |  |  |  |
| **Bile duct diameter, mean (SD), mm** | 10.8 (3.3) | 9.7 (3.2) | 12.4 (2.7) | **<0.001** |
| **Procedure duration\*, median (IQR), min** | 92.0 (71-110) | 82.5 (61-94) | 96 (90-120) | **0.006** |
| **EUS-assisted ERC type, n (%)** |  |  |  | **<0.001** |
| EUS-guided RV (guidewire) | 52 (75.4) | 24 (60) | 28 (96.5) |  |
| EUS-guided colorant injection (wireless RV) | 12 (17.4) | 12 (30) | 0 |  |
| Combined procedures | 5 (7.2) | 4 (10) | 1 (3.4) |  |
| **Needle size, n (%)** |  |  |  | **0.044** |
| 19 G | 34 (49.2) | 15 (37.5) | 19 (65.5) |  |
| 22 G | 31 (44.9) | 23 (57.5) | 8 (27.5) |  |
| Both needle sizes | 4 (5.8) | 2 (5.0) | 2 (6.9) |  |
| **Puncture site, n (%)** |  |  |  | **0.013** |
| Intrahepatic | 10 (14.5) | 3 (7.5) | 7 (24.13) |  |
| Extrahepatic | 57 (82.6) | 37 (92.5) | 20 (68.9) |  |
| Both | 2 (2.8) | 0 | 2 (6.9) |  |
| **Approach route, n (%)** |  |  |  | **0.003** |
| Transgastric | 12 (17.4) | 3 (7.5) | 9 (31) |  |
| Transduodenal: | 55 (79.7) | 37 (92.5) | 18 (62.0) |  |
| Both (gastric and duodenal) | 2 (2.89) | 0 | 2 (6.9) |  |
| **Scope-position (transduodenal)‡, n(%)** |  |  |  | **>0.99** |
| Short-scope position | 36 (66.0) | 23 (57.5) | 13 (44.8) |  |
| Long-scope position | 3 (5.5) | 2 (5.0) | 1 (3.4) |  |
| **Same session, n (%)** | 33 (47.8) | 10 (25.0) | 23 (79.3) | **<0.001** |
| **Other session, n (%)** | 36 (52.2) | 30 (75.0) | 6 (20.7) | **<0.001** |
| **Time from failed ERC, days, mean (SD)** | 11.9 (19.6) | 18.9 (23.0) | 2.14 (5.4) | **<0.001** |
| **EUS-FNA at same procedure, n (%)** | 14 (20.2) | - | 14 (48.2) | - |
| **Clinical outcomes, n (%) [95%CI]:** |  |  |  |  |
| **Overall technical success** | 55 (79.7) [68.3-88.4] | 36 (90.0) [76-97] | 19 (65.5) [46-82] | **0.028** |
| **Overall success** | 51 (73.9) [61.0-83.7] | 33 (82.5) [67-93] | 18 (62.1) [42-79] | 0.103 |
| **Overall adverse events rate** | 17 (24.6) [15.1-36.5] | 6 (15.0) [6-30] | 11 (37.9) [21-58] | 0.058 |
| **Related mortality** | 4 (5.7) | 1 (2.5) | 3 (10.3) | - |

Values are number (%) unless otherwise stated. \* Data missing for 12 patients. ‡ Data missing for 30 patients. ERC, endoscopic retrograde cholangiography; EUS, endoscopic ultrasound; FNA, fine needle aspiration; RV rendezvous.

### Global outcomes for EUS-assisted ERC

Technical and overall success rates of EUS-assisted ERC were 79.7% (95%CI, 68.3-88.4) and 74% (95%CI, 61.9-83.7), respectively.

Failed RV-cases were mainly related to guidewire manipulation (30%, **TableS1**). In supplementary **TableS2**, there is detailed information for each failed EUS-assisted ERC case.

### Analysis of malignant vs benign groups

Comparison of benign and malignant groups evidenced higher success for benign cases: a greater technical success of 90% (95%CI, 76-97) vs 65% (95%CI, 46-82), and overall success of 82% (95%CI, 67-93) vs 62% (95%CI, 42-79), (**Table 2, 3).**

Detailed information on EUS-assisted ERC technical success between malignant and benign groups is presented in **TableS1**.

### Safety

Seventeen AEs were detected (24.5%; 95%CI, 15.1-36.5), with a higher rate for malignant cohort vs benign: 37.9% (95%CI, 21-58) vs 15% (95%CI, 6-30) (**Table3**).

**Table 3:** Comparison of clinical outcomes between groups.

**Tabla 3:** Comparison of clinical outcomes between groups.
| EUS-guided assisted ERC: benign vs malignant |  |  |  |  |  |  |
| --- | --- | --- | --- | --- | --- | --- |
|  |  | No | Yes | OR [95%CI] | P value | N |
| Overall technical success, n (%): | Benign | 4 (10.0) | 36 (90.0) | Ref. | Ref. | 69 |
|  | Malignant | 10 (34.5) | 19 (65.5) | 0.21 [0.05-0.72] | **0.017** |  |
| Overall success, n (%): | Benign | 7 (17.5) | 33 (82.5) | Ref. | Ref. | 69 |
|  | Malignant | 11 (37.9) | 18 (62.1) | 0.35 [0.11-1.03] | 0.067 |  |
| Overall adverse events rate, n (%): | Benign | 34 (85.0) | 6 (15.0) | Ref. | Ref. | 69 |
|  | Malignant | 18 (62.1) | 11 (37.9) | 3.46 [1.13-11.54] | **0.036** |  |
| Comparison of outcomes between EUS-assisted ERC and EUS-transluminal groups, for malignant cases |  |  |  |  |  |  |
|  |  | No | Yes | OR [95%CI] | P value | N |
| Overall technical success, n (%): | Rendezvous | 10 (37.0) | 17 (63.0) | Ref. | Ref. | 104 |
|  | Transmural | 3 (3.90) | 74 (96.1) | 14.51 [3.96-70.13] | **<0.001** |  |
| Overall success, n (%): | Rendezvous | 11 (40.7) | 16 (59.3) | Ref. | Ref. | 104 |
|  | Transmural | 14 (18.2) | 63 (81.8) | 3.09 [1.18-8.16] | **0.026** |  |
| Overall adverse events rate, n (%): | Rendezvous | 16 (59.3) | 11 (40.7) | Ref. | Ref. | 104 |
|  | Transmural | 64 (83.1) | 13 (16.9) | 0.3 [0.11-0.78] | **0.017** |  |

ERC, endoscopic retrograde cholangiography; EUS, endoscopic ultrasound.

Most of the reported SAEs were perforation (n=5) or infection-related (n=5) and were detected as immediate or early-AEs (<7days). In the malignant group, most of the AEs where cholangitis and sepsis, likely related to failure to achieve adequate biliary drainage. All reported AEs, including severity (AGREE), timing, and type are detailed in **Supplementary TableS3**.

Related mortality was detected in 4 cases: 3 in the malignant group (2 perforation, 1 sepsis) vs. 1 in the benign group (1 pancreatitis).

### Univariate and multivariate analysis

On univariate and multivariate analysis, malignant pathology was statistically associated with a greater possibility of failure rate (technical success OR,0.21; 95%CI, 0.05-0.72; p=0.017), and higher risk for AEs (OR,3.46; 95%CI, 1.13-11-5; p=0.036) for EUS-RV group. Concerning overall success, a non-significant trend was observed (OR,0.35; 95%CI, 0.11-1.03; p=0.067) (**Table 2, 3 and FigureS4**).

No statistically significant differences in terms of puncture site, approach route, needle type, rendezvous type, anticoagulation, or albumin level were related to success or safety. Complete statistical results can be found in **Supplementary material (Table S4, S5).**

### Comparative analysis with EUS-TMD group for malignant disorders

The EUS-TMD group had significantly greater bile duct diameter (mean [SD]: 16.8 [3.9] vs 12.7 [2.6] mm, p<0.001) and higher Charlson comorbidity index (11[1.5] vs 9.46 [2.8], p=0.028) compared to the malignant EUS-RV group. Technical success, overall success, and adverse events for EUS-TMD were 96% (95%CI, 89-99), 81.8% (95%CI, 71-90), and 16.9% (95%CI, 9-27) respectively (**Table4**).

**Table 4:**
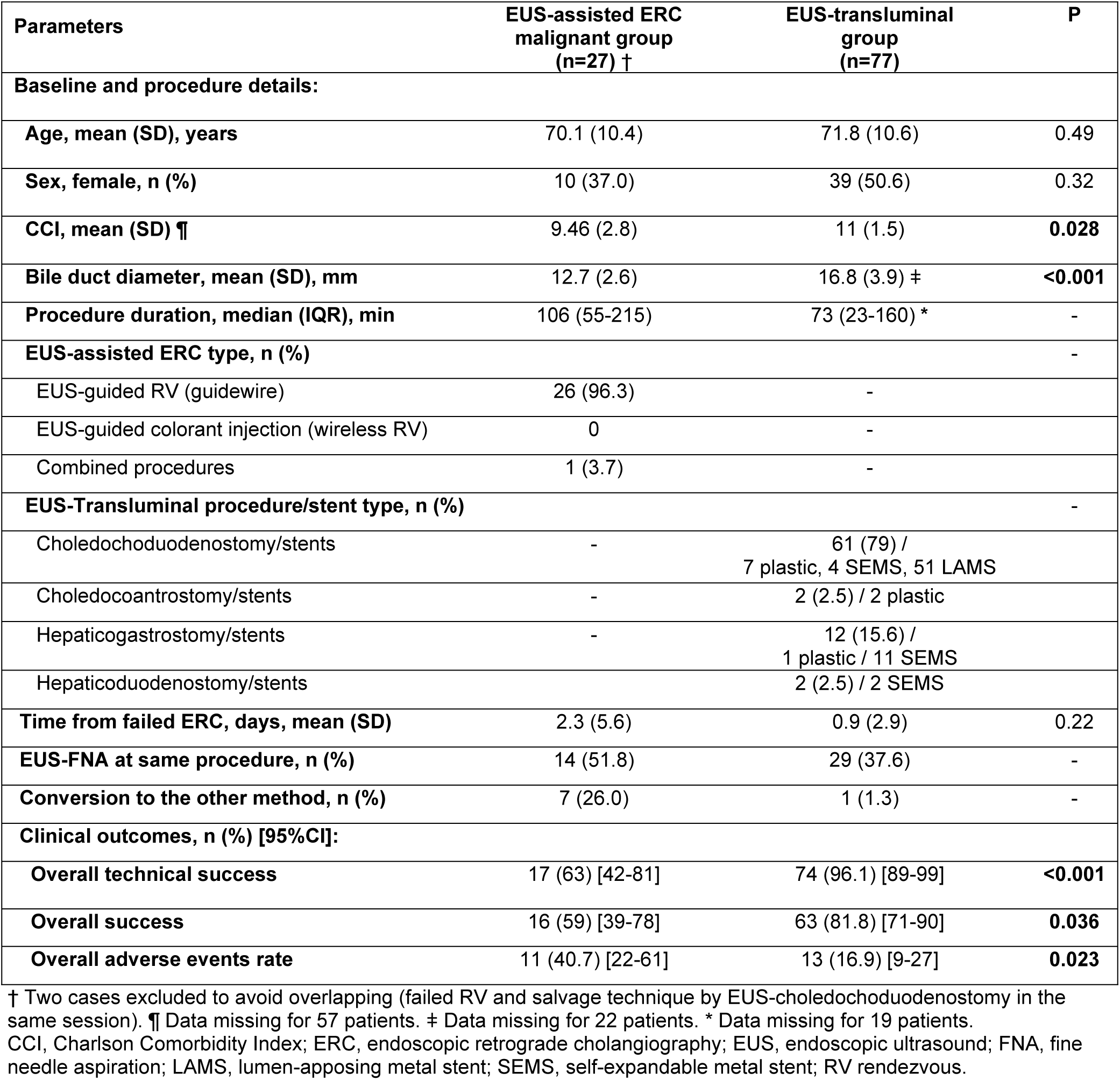
Comparison of baseline characteristics, procedure details, and outcomes between EUS-assisted ERC and EUS-transluminal groups, for malignant cases.

Concerning outcomes comparison, better results were encountered for the EUS-TMD group: greater technical success (OR, 14.51; 95%CI, 3.96-70.13; p<0.001) and overall success (OR,3.09; 95%CI, 1.18-8.16; p=0.026), and lower AEs rate (OR, 0.30; 95%CI, 0.11-0.78; p=0.017) **(Table3 and FigureS5).**

It must be noted that seven failed EUS-RV were successfully rescued with EUS-TMD procedures, and only one failed transluminal drainage (choledochoduodenostomy) was rescued with RV [9].

## DISCUSSION

This is a retrospective study of patients from a single tertiary referral center who underwent EUS-assisted-ERC after unsuccessful ERC over an 11-year period. The number of EUS-RV (n=69, 0.76%) and or EUS-guided biliopancreatic intervention (n-183, 1.7%) represents a low volume in a high-volume center, with only 6.2 EUS-guided biliary RV cases annually. So, this serves to highlight the efficacy of standard ERC.

The EUS-guided RV technique is a technically demanding alternative technique and not free from AEs, due to several technical factors, as well as the lack of dedicated accessories [1,10]. Previous studies have reported that EUS-RV seems to be feasible after failed ERC, but there is wide variability in the reported success and AEs rates [2,11]. Although EUS-RV is one of the most widely practiced techniques, in our opinion, and in accordance with guidelines and reviews, EUS-RV should be considered after a failed ERC in centers where expertise is available, and preferably during the same session [1-3,11].

In our study, the technical success rate was 79% and the AEs rate was 24%. **TableS6** contains a literature review of most EUS-RV studies compared with our study [12-29]. It includes 585 cases (19 studies), with a technical success rate of 85.4%, and AEs rate of 15.5%. The lack of standardization makes comparison of results difficult, where heterogeneity is noted. Recently, Yoon et al. summarized the outcomes of 525 cases in a meta-analysis, with most of the published series including only a small number of cases. The pooled rate for technical success was 88% (benign/malignant, 89/90%) and for overall AEs 14% [11].

This current study included a greater number of patients, and the results were comparable to although more modest those revealed in these recent meta-analyses [2,11]. Rigorous data collection in prospective databases from a single-center (specially for AEs), a long study period including all consecutive cases from introduction of the procedure, and inclusion of a malignant group (predominantly palliative cases) may partially explain these differences. Regarding patients with normal anatomy vs surgically altered anatomy, this study includes only cases with accessible papilla, in accordance with dedicated literature suggesting that PTBD and EUS-transmural BD seem to be better suited to this scenario [1,3,11].

EUS-RV is an example of an assisted or ‘indirect’ technique in which EUS facilitates the introduction of a guidewire towards the papilla to within reach of a duodenoscope without involving tract dilation or stent transmural placement. In addition, although the procedure may sound simple, it is a multiple-step process and can be challenging even for an experienced endoscopist [30]. The most challenging points during EUS-RV are: i) guidewire manipulation through a rigid long needle into the bile duct and across the stenosis/papilla with limited directional manoeuvrability, and limited needle angulation; ii) a complex endoscope exchange process that may cause guidewire loss; and iii) retrograde biliary cannulation (alongside or over-the-wire) [2,14]. In this study, in accordance with prior experience, the main cause of failed EUS-RV was the inability to advance the guidewire through the papilla. But in 3 cases a guidewire dislodgement during scope exchange was also noted (**TableS2**), altering the overall success rate [10].

Concerning recommended approach routes (intra- or extrahepatic) and scope-positions (long or short), no robust data exists to recommend one over the other. Recent literature shows extrahepatic and short scope position as preferred because the shorter distance (biliary access point and obstruction) allows for better manoeuvrability while advancing the guidewire, while greater diameter of CBD allows for easier targeting. Short-scope position is superior to long-scope because a duodenal bulb with long-position will obligate the guidewire advance upstream in the direction of the liver rather than to the distal CBD [11,30,31]. Similarly, extrahepatic/transduodenal route and short-scope position were more common in our study, in both groups. Although bile leak from the punctured biliary duct has been described as a potential AE with the extrahepatic approach, no cases were detected in our study [14,31].

According to a recent review, EUS-RV has the potential advantage of fewer AEs when compared with PTBD, percutaneous-RV, and EUS-BD [11,14]. In our study, the AEs rate and mortality were strikingly high in the malignant group, especially compared to the benign diseases group and the EUS-TMD technique. Most of the AEs reported in the RV-malignant group where cholangitis or sepsis, likely related to failure to achieve adequate biliary drainage. Also, a longer procedure time during the same failed ERC session, with a more demanding tight tumoral stenosis, high CCI index, and fragility for this malignant group, may explain these findings. In contrast, AEs for the RV/benign and EUS-TMD groups were similar to what is reported in the literature.

With the goal of reducing the cholangitis rate, after gaining more experience with this demanding procedure, cholangiogram (contrast injection) was progressively avoided without increasing the failure rate. In this manner, after a failed RV, the level of concern about an urgent PTBD was more acceptable.

Current international guidelines recommend RV techniques in benign diseases but they can also be used in malignant biliary diseases [1,11]. In particular, the latest ESGE guidelines state that EUS-RV should be the first-line approach following failed ERCP, over PTBD or EUS-BD, in benign diseases if expertise is available. Comparing benign vs malignant cohorts, the technical success of EUS-RV in benign disease may be lower than in malignant disease, and AEs may be more likely to occur, owing to limited bile duct dilatation [1]. Interestingly, in our study, a small bile duct diameter was associated with the benign group, but shorter procedure time and a significantly higher technical success with a lower AEs rate was seen.

Our initially standard approach to EUS-guided biliary access for malignant distal biliary obstruction changed gradually from EUS-RV to EUS-biliary TMD. This decision was based on the progressive perception of shorter procedure time, superior technical success, lower AEs rates, and successful salvage transmural drainage after failed RV following the introduction of biliary LAMS at our unit in 2014 [8]. This attractive and effective EUS-transmural BD variant has gained greater prominence in malignant distal obstructions. This study provides a direct comparison between the two techniques, clearly favouring the EUS-TMD group (basically, choledochoduodenostomy, hepaticogastrostomy) over EUS-RV with greater success, shorter procedure duration, and better safety rates.

Thus, with our findings, we feel that EUS-transmural BD drainage may remain as the preferred approach over EUS-RV in malignant diseases. EUS-RV might be reserved for benign disease.

This study has some limitations, mainly owing to its retrospective design. First, because of a lack of data, patient discomfort, hospitalization, and cost comparison could not be assessed. Second, the variability of assisted techniques (colorant/guidewire) included in this study may have imposed a selection bias on the study population. Third, this technique may cause AEs related to the ERC or EUS-guided biliary access, but it is difficult to ensure causality; thus an overestimation of AEs related to EUS may have occurred. For this reason, two cases (EUS-RV and EUS-TMD in the same procedure) were excluded from the EUS-RV group for the comparison analysis with EUS-TMD. Fourth, this single-center study may not be applicable to other centers with different practice patterns. Lastly, the lack of a standardized or uniform protocol due to changes related to the long study period, with no specific follow-up, may have entailed a lack of some relevant data. Furthermore, comparison of RV and TMD was limited because of the non-randomized nature of the study and the step-up approach in which both techniques were used.

Among the strengths of this study are its rigorous design and analysis of specific and detailed data, inclusion of benign and malignant diseases, and consecutive inclusion during a long period with a single operator in a tertiary center, providing greater homogeneity in the results.

This study offers new and relevant information, not available in the latest guidelines or reviews. The EUS-RV technique is a technically demanding multi-step technique, with better effectiveness in benign than in malignant disorders. In case of failed ERC and accessible papilla, when comparing EUS-RV with EUS-biliary TMD in malignant diseases, EUS-RV has a lower success rate, higher AEs, and a worrying mortality rate.

Therefore, given these findings, maybe EUS-RV is not the best option for malignant disorders, and might be reserved for benign cases. Innovations with new devices are needed to simplify and improve the range of EUS-guided interventions.

## Data Availability

All relevant data are within the manuscript and its Supporting Information files.

NA

## Acronyms

AE: adverse event
CBD: common bile duct
ERCP: endoscopic retrograde cholangiopancreatography
EUS: endoscopic ultrasound
LAMS: lumen apposing metal stent
MBO: malignant biliary obstruction
RV: rendezvous

